# Do Large Language Models Use the Clinical Vignette? A Question-Ablation Study on the Orthopaedic In-Training Examination

**DOI:** 10.64898/2026.09.15.26363139

**Authors:** Fatima Gafoor, Ma’az Syed, Mansur M. Halai, Nicholas J. Yee

## Abstract

**Background:** Large language models (LLMs) have demonstrated strong performance on standardized medical examinations, with recent studies reporting performance approaching or exceeding that of senior medical residents. However, examination accuracy alone does not establish how models arrive at their answers or the relative contributions of the clinical vignette, imaging, and answer options to model performance. We evaluated the contribution of these question components to LLM performance on the Orthopaedic In-Training Examination (OITE).

**Methods:** We evaluated three open-source Ministral-3 models (3B, 8B, and 14B) and five proprietary models (Claude Haiku-4.5, Sonnet-4.6, Opus-4.8, GPT-5.6 Luna, and GPT-5.6 Terra) using 792 OITE questions from 2020 through 2024, comprising 434 questions containing clinical images and 358 without images. Question components were sequentially removed across five image-containing and three non–image-containing conditions. Accuracy was summarized using the median and IQR and compared using paired analyses with Holm adjustment. Semantic similarity between model-generated explanations and reference discussions was assessed using BioMedBERT.

**Results:** For image-containing questions, pooled accuracy was 59.13% (58.21–60.08) with complete information and 59.13% (58.18-60.11) without images. Accuracy decreased to 49.05% (48.07– 50.00) without the clinical vignette, 45.68% (44.70–46.69) without the vignette and images, and 36.49% (35.51–37.44) when the vignette, images, and question were removed. For non-image-containing questions, accuracy decreased from 72.94% (72.10–73.85) with complete clinical context to 53.53% (52.41–54.68) without the vignette and 37.36% (36.28–38.48) with answer options alone. OpenAI GPT-5.6 Terra achieved the highest full-context accuracy in both image-containing 81.80% (80.41–82.95) and non-image-containing 94.13% (93.30–94.97) questions. Removal of images alone did not significantly affect accuracy for any model, whereas removal of the clinical vignette significantly reduced accuracy for most models. When provided with only the answer options, without the question stem, clinical context, or images, the observed accuracy for every model exceeded the 25% expected from random selection. Specifically, the highest accuracy was 45.62% (44.24–47.24) for Claude Opus 4.8 on questions originally containing images and 46.09% (44.41–47.77) for GPT-5.6 Terra on questions without images. Although closed-source models achieved substantially higher accuracy than open-source models, all models performed above chance when provided answer options alone. BioMedBERT similarity remained high despite substantial differences in accuracy and was not significantly associated with accuracy by Spearman correlation (ρ=−0.17; *P*=0.29).

**Conclusion:** Clinical vignettes were the primary contributor to improving LLM performance on OITE whereas images had minimal effect. The high accuracy when only answer options were given suggests the models may be learning spurious correlations. Furthermore, BioMedBERT semantic similarity remained high across models despite wide differences in accuracy and did not consistently distinguish correct from incorrect responses. Thus, high examination performance may not directly reflect clinical reasoning, highlighting the importance of assessing how LLMs use the information provided when interpreting benchmark performance.

## INTRODUCTION

The Orthopaedic In-Training Examination (OITE) is an annual, multiple-choice assessment developed by the American Academy of Orthopaedic Surgeons (AAOS, Rosemont, USA) to support knowledge assessment of orthopaedic surgery residents across all training levels. Residency programs use OITE results to identify learning needs and evaluate educational outcomes, while residents may use the examination as a longitudinal benchmark of progress. The examination spans major topics of orthopaedic surgery and may include clinical vignettes and imaging.

Rapid advances in large language models (LLMs), artificial intelligence systems that interpret and generate human-like language, have introduced new possibilities for medical education, clinical decision support, and knowledge assessment. Researchers have systematically evaluated LLMs on a wide range of standardized medical licensing and specialty board examinations and performance.^1,2^ Within orthopaedic surgery, several studies have evaluated LLM performance on the OITE. Lubitz and Latario compared ChatGPT and Google Bard on the 2022 OITE, while Xu et al. subsequently demonstrated significantly higher (*P*<0.01) performance with GPT-4 (74.1%) than GPT3.5 (50.3%) and Bard (58.2%) on the same exam.^3,4^ More recent studies have reported further improvements with newer model generations, with contemporary models approaching or exceeding the performance of senior orthopaedic residents.^5,6^ Additional work has evaluated LLMs on orthopaedic board-style questions in formats beyond the OITE.^7^

Raw examination accuracy, however, does not establish how a model has arrived at an answer. A model may use the clinical vignette, interpret accompanying images, recognize patterns in the answer choices, or reproduce material encountered during LLM training. Prior orthopaedic benchmarks have generally tested a single prompt configuration on one or a small number of closed-source proprietary systems that do not share details on their model architecture or training data. These experimental designs cannot isolate the marginal value of clinical context or images, and they provide limited information about performance at different LLM parameter sizes.

The primary aim of this study was to characterize the relative contributions of clinical context, associated images, and questioning to LLM performance on the OITE by systematically varying the information provided to each model. Secondary aims were to compare the performance of open-source versus closed-source proprietary LLMs and to examine the relationship between model scale and examination accuracy. Understanding the components affecting LLM performance and assessing the quality of its answer explanations are important for understanding its clinical reasoning and to build clinicians’ trust in the systems. We evaluated three open-source models from the Ministral-3 family (3-Billion, 8-Billion, and 14-Billion parameter variants) and five proprietary frontier models, including Claude- and OpenAI GPT-based systems, using OITE questions from five consecutive examination years from 2020 through 2024. We hypothesized that clinical context would be the primary contributor to model performance, whereas associated radiological and clinical images would provide a comparatively smaller incremental benefit to performance. We further hypothesized that proprietary models would outperform the evaluated open-source models and that larger models would demonstrate greater examination accuracy.

## METHODS

### Study Design and Data Source

We conducted a retrospective benchmarking study using question content from the OITE 2020 through 2024 examination years. The study was designed to determine how LLM performance changes according to the amount and type of information provided. Specifically, we evaluated the contributions of the clinical vignette, final question sentence, answer options, and accompanying images by presenting the same questions under different information conditions. Each question and experimental condition were evaluated in an independent model session without prior conversation history, question-specific examples, or topic-specific priming. Models received no additional fine-tuning beyond their existing training and the standardized study prompts.

The question items contained the clinical vignette, the image references where applicable, the final question sentence, the complete set of answer options, and the correct answer for each question. Questions without a separable clinical vignette (i.e., questions consisting only of a single question sentence) were excluded because removal of the clinical vignette would not produce a distinct experimental condition.

The initial data set contained 1051 questions. After excluding 259 questions without a clinical vignette, 792 distinct questions were included in the final analysis. Questions were categorized according to the presence of clinical images in the original examination question. Image-containing questions (ICQs) comprised 434 questions that included one or more clinical images, whereas non-image-containing questions (NICQs) comprised 358 questions without associated clinical images. Images were presented in their original format, with radiographs in grayscale and non-radiographic clinical images in color. ICQs were evaluated under five conditions that varied in the offering of the clinical stem, question sentence, answer options, and images (Table 1). NICQs were evaluated under three conditions because comparisons with and without images were not applicable (Table 2).

**Table 1.** Representative examples of the five experimental conditions applied to the OITE image-containing questions containing clinical images.

| Experimental Conditions (ICQ) | Example |
| --- | --- |
| Full question stem with images (baseline) | Figures 1 and 2 are the radiographs of a 61-year-old patient 5 and a half months after surgical intervention for distal tibia fracture. The patient has a 40-year history of tobacco use.<br>[ Figure 1 image ]<br>[ Figure 2 image ]<br>If this issue persists at their one-year follow-up, what surgical approach would be best for revision?<br>A) Anterolateral, B) Medial, C) Posterolateral, D) Anterior |
| Without images (W/O<sub>images</sub>) | Figures 1 and 2 are the radiographs of a 61-year-old patient 5 and a half months after surgical intervention for distal tibia fracture. The patient has a 40-year history of tobacco use.<br>If this issue persists at their one-year follow-up, what surgical approach would be best for revision?<br>A) Anterolateral, B) Medial, C) Posterolateral, D) Anterior |
| Without stem (W/O<sub>stem</sub>) | [ Figure 1 image ]<br>[ Figure 2 image ]<br>If this issue persists at their one-year follow-up, what surgical approach would be best for revision?<br>A) Anterolateral, B) Medial, C) Posterolateral, D) Anterior |
| Without stem and without images (W/O<sub>stem+images</sub>) | If this issue persists at their one-year follow-up, what surgical approach would be best for revision?<br>A) Anterolateral, B) Medial, C) Posterolateral, D) Anterior |
| Without stem, images, and question (W/O<sub>stem+images+question</sub>) | A) Anterolateral, B) Medial, C) Posterolateral, D) Anterior |

ICQ denotes image-containing question; OITE, Orthopaedic In-Training Examination; and W/O, without.

**Table 2.** Representative examples of the three experimental conditions applied to OITE questions without associated clinical images.

| **Experimental Conditions (NICQ)** | **Example** |
| --- | --- |
| Full question stem | A 27-year-old man has recalcitrant left shoulder pain. On examination, he has weakness of flexion at 90° of abduction and weakness of external rotation with the arm at the side.<br>What is the most common location at which a compressive neuropathy would result in these signs and symptoms?<br>A) Suprascapular notch, B) Spinoglenoid notch, C) Quadrilateral space, D) C2-C3 foramen |
| Without stem | What is the most common location at which a compressive neuropathy would result in these signs and symptoms?<br>A) Suprascapular notch, B) Spinoglenoid notch, C) Quadrilateral space, D) C2-C3 foramen |
| Without stem and question | A) Suprascapular notch, B) Spinoglenoid notch, C) Quadrilateral space, D) C2-C3 foramen |

NICQ denotes non–image-containing question; OITE, Orthopaedic In-Training Examination; and W/O, without.

### Models and Deployment

Eight LLMs were evaluated. The open-source model group comprised three models from the Ministral-3 family: Ministral-3 3B, Ministral-3 8B, and Ministral-3 14B (which were pretrained from the March 2025 Mistral Small 3.1 model, which has no publicly disclosed knowledge cutoff date; Mistral AI SAS, Paris, France). The proprietary model group included the Claude family (Haiku 4.5, Sonnet 4.6, Opus 4.8; Anthropic PBC, San Francisco, USA), with knowledge cutoff dates of February 2025, May 2025, and January 2026, respectively. The OpenAI family included GPT-5.6 Luna and GPT-5.6 Terra (OpenAI, San Francisco, USA), both with a knowledge cutoff date of February 16, 2026. All models were evaluated using the same general prompting framework. The Ministral-3 models were deployed locally through Ollama version 0.30.10 on a workstation equipped with two NVIDIA GeForce RTX 5060 Ti GPU (16GB each; Nvidia Corporation, Santa Clara, USA). Proprietary models were accessed through the Claude and OpenAI API, respectively.

### Prompt Design and Output Format

A standardized system prompt was used across all models and experimental conditions (Table S1). Models were instructed: “You are an expert orthopaedic surgery exam assistant” and were directed to answer questions using the information and multiple-choice options provided. The system prompt required models to select only from the supplied answer options and to provide a response even when information was incomplete or unavailable.

The user-prompt template was defined in a YAML configuration file and standardized the presentation of each question across models and experimental conditions. Each item was separated into three components: the clinical vignette, the final question sentence, and the answer options. These components and any accompanying images were included or removed according to the assigned condition (ICQ vs NICQ).

For each question, models were instructed to carefully evaluate the information provided and select exactly one answer option. Responses were restricted to a structured JSON format containing two fields, the answer and the reasoning. Semantic similarity between LLM-generated explanations and the reference discussion for each question was quantified using BioMedBERT. ^8^ Models received no feedback regarding whether their responses were correct.

### Experimental Conditions

The included OITE questions were evaluated under different information conditions based on whether the clinical stem, final question sentence, answer options and images were provided to the model.

ICQs were tested under five conditions (Table 1). The baseline (ICQ_baseline_) contained all available information: the clinical stem, final question sentence, answer options, and images. The experimental conditions were tested by removing only the images (ICQ-W/O_images_), removing only the stem (ICQ-W/O_stem_), removing both the stem and images (ICQ-W/O_stem+images_), and removing everything except the answers (ICQ-W/O_stem+images+question_).

NICQs were tested under three conditions (Table 2). The baseline contained all available information: the clinical stem, final question sentence, and answer options (NICQ_baseline_). The experimental conditions were tested by removing the clinical stem (NICQ-W/O_stem_) and by removing everything except the answer options (NICQ-W/O_stem+question_).

### Data and Statistical Analysis

Accuracy was calculated as the percentage of questions answered correctly and was reported separately for each model and experimental condition for ICQs and NICQs. Overall accuracy across models was also calculated for each OITE exam year. Performance in W/O_images+stem+question_ was compared with the expected accuracy of 25% from random selection among four answer options.

For each model and information condition, question-level outcomes were resampled with replacement 1,000 times. Each resample contained the same number of questions as the original sample. Results were reported as the median accuracy across the bootstrap samples, with the 25th and 75th percentiles reported as the bootstrap interquartile range (IQR). For model-specific comparisons between experimental conditions, paired binary outcomes for the same questions were compared using McNemar tests with Holm adjustment ( 0.05). A Spearman’s rank correlation was used to assess the association between answer accuracy and the semantic similarity of LLM-generated explanations to the reference OITE explanations, as measured by BioMedBERT.

## RESULTS

### Image-Containing Questions

Across all evaluated models, the overall accuracy, defined as the proportion of model responses matching the reference answer provided for each OITE question, was 59.13% was 59.13% (IQR, 58.21-60.08) when the complete clinical vignette, question sentence, answer options and associated images were provided (ICQ_baseline_). Removal of the clinical images while preserving the complete context (ICQ-W/O_images_) resulted in the same overall accuracy of 59.13% (IQR, 58.18–60.11). In contrast, the removal of the clinical vignette when images remained available (ICQ-W/O_stem_) reduced performance to 49.05% (IQR, 48.07-50.00). When both the vignette and images were removed (ICQ-W/O_images+stem_), there was a further reduction to 45.68% (IQR, 44.70-46.69). Performance was lowest when the models received only the answer options (ICQ-W/O_images+stem+question_), with an overall accuracy across models of 36.49% (IQR, 35.51-37.44; Table 3).

**Table 3.**
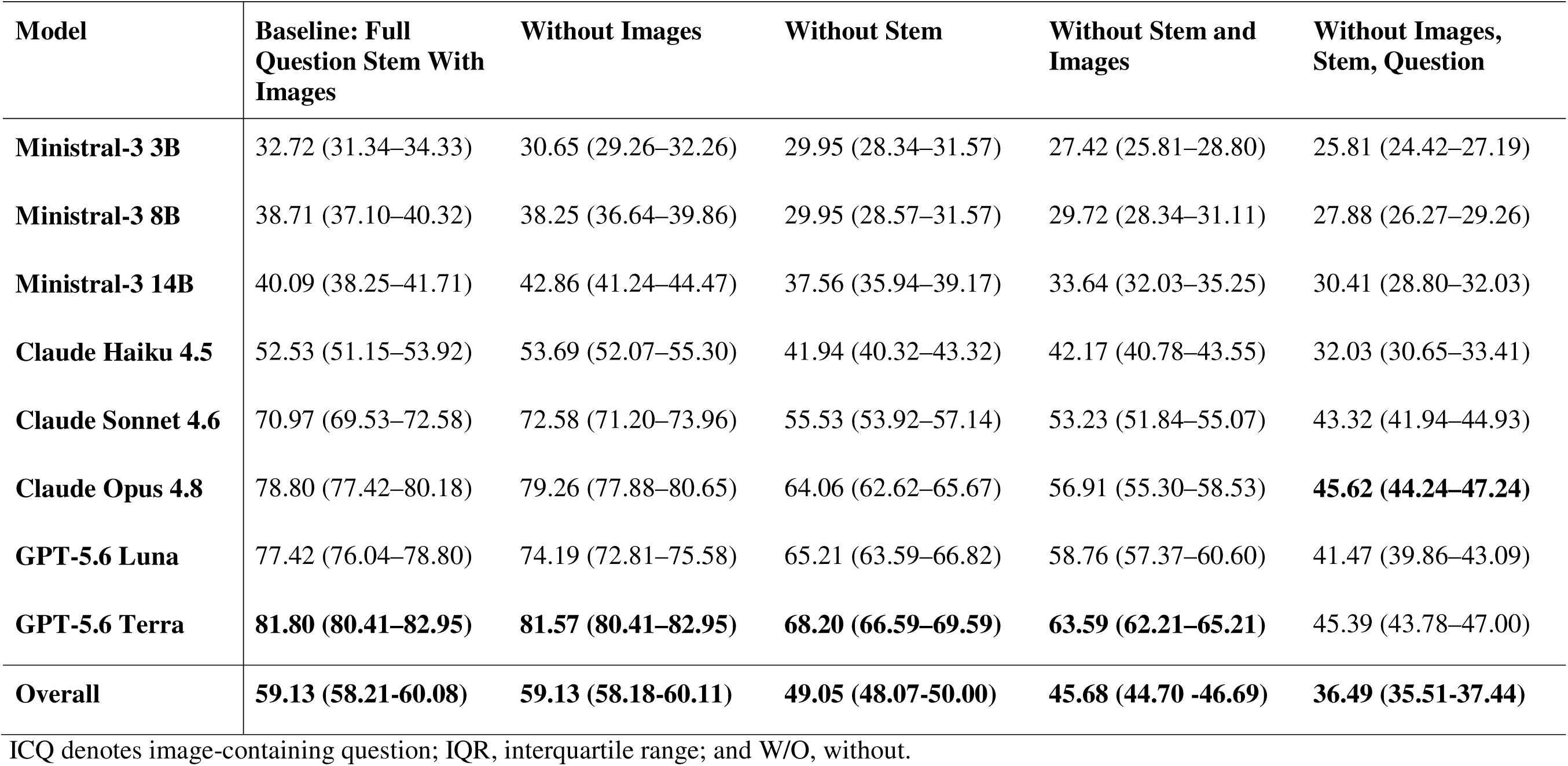
ICQs Accuracy (%) of large language models on OITE questions containing clinical images across five experimental conditions. Bootstrap interquartile ranges (IQRs) are shown in parentheses.

Among the open-source models, Ministral-3 14B achieved the highest accuracy under the full-information condition (ICQ_baseline_), at 40.09% (IQR, 38.25–41.71), followed by Ministral-3 8B at 38.71% (IQR, 37.10–40.32) and Ministral-3 3B at 32.72% (IQR, 31.34–34.33). Closed-source proprietary models demonstrated higher accuracy, with GPT-5.6 Terra achieving the highest performance under the full-information condition at 81.80% (IQR, 80.41–82.95), followed by Claude Opus 4.8 at 78.80% (IQR, 77.42-80.18), GPT-5.6 Luna at 77.42% (IQR, 76.04–78.80), Claude Sonnet 4.6 at 70.97% (IQR, 69.53-72.58), and Claude Haiku 4.5 at 52.53% (IQR, 51.15–53.92). Withholding clinical images while keeping the complete clinical context (W/O_images_) resulted in minimal changes in model performance compared to the full-information condition (ICQ_baseline_), with absolute differences ranging from 0.23 to 3.23 percentage points, across all the evaluated models. Accuracy generally declined as the clinical information was incrementally removed, although closed-source proprietary models consistently maintained higher performance than open-source models across all experimental conditions.

Removal of the clinical vignette significantly reduced accuracy in 7 of 8 models (adjusted (all *P* 0.02); Table S2), with reductions ranging from 8.76 to 15.44 percentage points. No significant difference was observed for Ministral-3 3B. In contrast, removing images alone did not significantly affect accuracy for any model (*P*=0.07-1.00). Changes in accuracy were small and ranged from a decrease of 3.23 percentage points for GPT-5.6 Luna to an increase of 2.77 percentage points for Ministral-3 14B.

Accuracy declined further as additional clinical information was removed, with the lowest accuracy observed when only answer options were provided (ICQ-W/O_images+stem+question_), with an overall decrease of 22.64 percentage points relative to baseline. Significant decreases in accuracy compared to baseline were observed for Ministral-3 8B, Ministral-3 14B, Claude Haiku 4.5, Claude Sonnet 4.6, Claude Opus 4.8, GPT-5.6 Luna, and GPT-5.6 Terra. Among these models, reductions ranged from 9.68 to 36.41 percentage points, adjusted *P* values were ≤0.004. The 6.91-percentage-point reduction for Ministral-3 3B was not significant (*P*=0.09; Table S2).

### Non-Image-Containing Questions

For non-image-containing questions, overall accuracy was highest under the full-information condition (NICQ-baseline; 72.94%; IQR, 72.10–73.85). Removal of the clinical vignette (NICQ-W/O_stem_) reduced overall accuracy to 53.53% (IQR, 52.41–54.68), with a further decline to 37.36% (IQR, 36.28– 38.48) when only the answer options were provided (NICQ-W/O_stem+question_; Table 4).

**Table 4.** NICQs Accuracy (%) of large language models on OITE questions without clinical images across three experimental conditions. Bootstrap interquartile ranges (IQRs) are shown in parentheses.

| **Model** | **Full Question Stem** | **Without Stem** | **Without stem and question** |
| --- | --- | --- | --- |
| **Ministral-3 3B** | 41.06 (39.39–42.74) | 34.08 (32.40–36.03) | 27.09 (25.42–29.05) |
| **Ministral-3 8B** | 54.75 (53.07–56.70) | 41.06 (39.39–43.02) | 31.01 (29.33–32.68) |
| **Ministral-3 14B** | 53.91 (51.96–55.87) | 40.78 (39.11–42.74) | 29.05 (27.65–30.73) |
| **Claude Haiku 4.5** | 70.67 (68.99–72.35) | 48.32 (46.65–50.28) | 32.12 (30.45–33.80) |
| **Claude Sonnet 4.6** | 87.15 (86.03–88.55) | 62.29 (60.34–63.97) | 44.97 (43.30–46.65) |
| **Claude Opus 4.8** | 89.94 (88.83–91.06) | 65.92 (64.25–67.60) | 45.25 (43.30–46.93) |
| **GPT-5.6 Luna** | 91.90 (90.78–92.74) | 66.76 (65.08–68.44) | 43.30 (41.34–44.97) |
| **GPT-5.6 Terra** | **94.13 (93.30–94.97)** | **68.99 (67.60–70.67)** | **46.09 (44.41–47.77)** |
| **Overall** | **72.94 (72.10–73.85)** | **53.53 (52.41–54.68)** | **37.36 (36.28–38.48)** |

IQR denotes interquartile range; NICQ, non–image-containing question; and W/O, without.

Among the open-source models, Ministral-3 8B achieved the highest baseline accuracy (54.75%; IQR, 53.07–56.70), followed closely by Ministral-3 14B (53.91%; IQR, 51.96–55.87) and Ministral-3 3B (41.06%; IQR, 39.39–42.74). Among the proprietary models, GPT-5.6 Terra achieved the highest baseline accuracy (94.13%; IQR, 93.30–94.97), followed by GPT-5.6 Luna (91.90%; IQR, 90.78–92.74) and Claude Opus 4.8 (89.94%; IQR, 88.83–91.06).

Under the answer-options-only condition (NICQ-W/O_stem+question_), all models achieved accuracies above the 25% expected by chance. Accuracy was 27.09% (IQR, 25.42–29.05) for Ministral-3 3B, 31.01% (IQR, 29.33–32.68) for Ministral-3 8B, and 29.05% (IQR, 27.65–30.73) for Ministral-3 14B. Among the closed-source proprietary models, accuracy was 32.12% (IQR, 30.45–33.80) for Claude Haiku 4.5, 44.97% (IQR, 43.30–46.65) for Claude Sonnet 4.6, 45.25% (IQR, 43.30–46.93) for Claude Opus 4.8, 43.30% (IQR, 41.34–44.97) for GPT-5.6 Luna, and 46.09% (IQR, 44.41–47.77) for GPT-5.6 Terra. GPT-5.6 Terra was the highest-performing model under this condition.

As with ICQs, model-specific paired analyses demonstrated significant reductions in accuracy for all eight models across all three NICQ comparisons: baseline versus NICQ-W/O_stem_, baseline versus NICQ-W/O_stem+question_, and NICQ-W/O_stem_ versus NICQ-W/O_stem+question_ (all *P*≤0.01; Table S3). From baseline to the answer-options-only condition, overall accuracy decreased by 35.58 percentage points, while model-specific reductions ranged from 13.97 to 48.60 percentage points (all adjusted P <0.001).

### BiomedBERT Results

Despite substantial differences in accuracy between models, median BioMedBERT similarity remained within a narrow range. For ICQ questions under the full-context condition (baseline), accuracy ranged from 32.72% to 81.80%, whereas median BioMedBERT similarity ranged from only 0.9893 to 0.9932. Although similarity increased modestly across the Ministral models, this pattern was not consistent across model families. Notably, GPT-5.6 Terra achieved the highest accuracy (81.80%) but had lower median similarity (0.9894) than Ministral-3 3B, which achieved the lowest accuracy (32.72%; similarity, 0.9927). A similar pattern was observed for NICQs, where accuracy ranged from 41.06% to 94.13% while median similarity remained between 0.9887 and 0.9935 (Table S4).

Similarity scores were also comparable between correct and incorrect responses across experimental conditions. For ICQs, significant differences were observed under the full-information (baseline) condition (median difference, −0.00049; *P*<0.001), after removal of the clinical vignette and images (ICQ-W/O_images+stem:_ +0.00019; *P*=0.02), and when only the answer options were provided (ICQ-W/O_images+stem+question_: +0.00117; *P*<0.001). For NICQs, a significant difference was observed after removal of the clinical vignette (NICQ-W/O_stem_: +0.00092; *P*<0.001), and when only the answer options were provided (NICQ-W/O_stem+question_: median difference, +0.00162; adjusted *P*<0.001). Absolute differences in median similarity ranged from 0.00002 to 0.00117 for ICQs and from 0.00016 to 0.00162 for NICQs. Although these comparisons reached statistical significance, the differences were small and were not consistent in direction. Detailed comparisons are reported in Table 5. Furthermore, when image-containing and non-image-containing questions were combined by model and OITE examination year, BioMedBERT similarity was not significantly associated with accuracy by Spearman rank correlation (ρ=-0.17, *P*=0.29; Figure 1).

**Figure 1.**
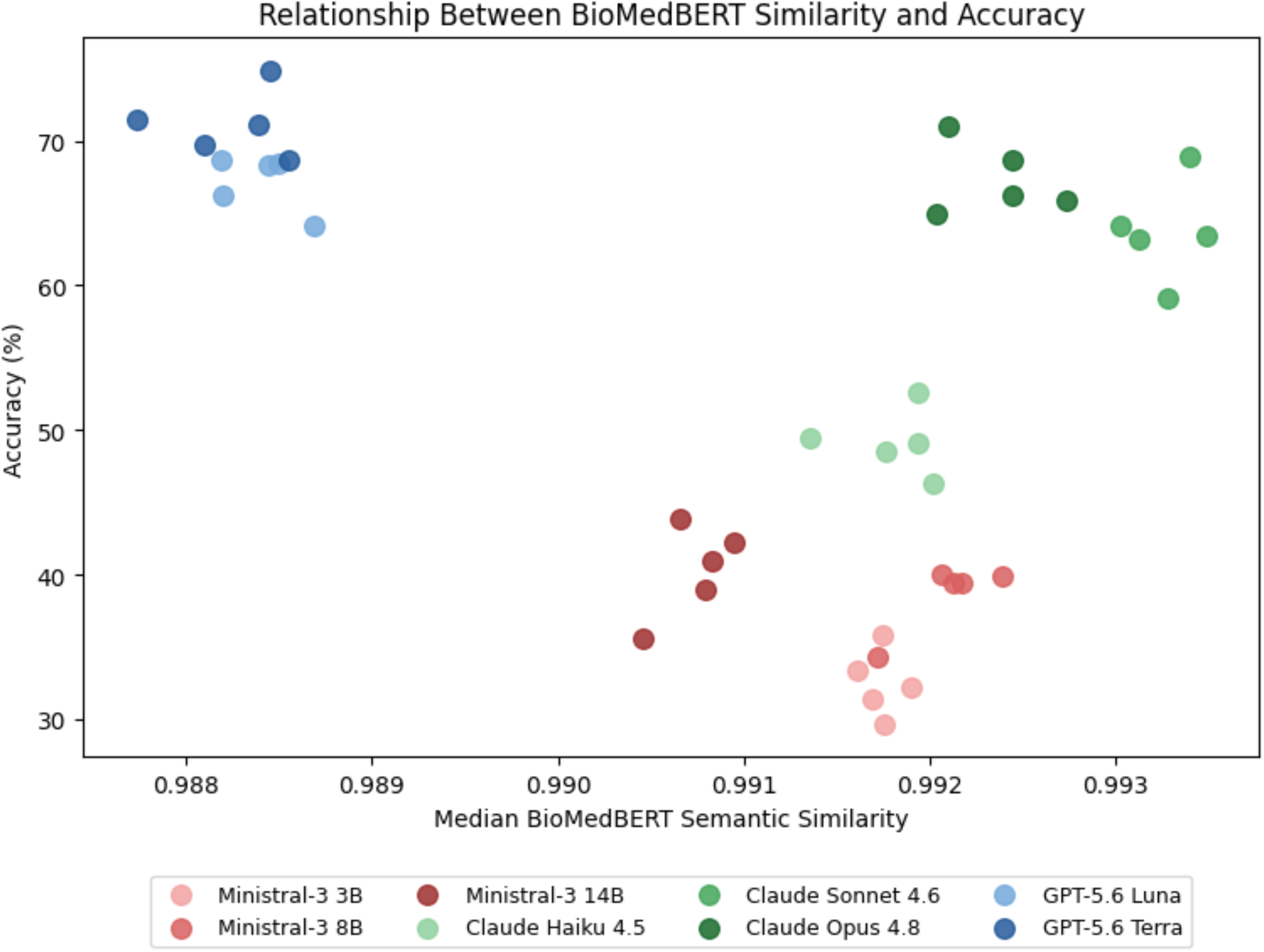
Relationship between BioMedBERT semantic similarity and answer accuracy across LLMs and each year of the OITE exam. Each point represents one model-examination year observation, with image-containing and non-image-containing questions combined.

**Table 5.** BioMedBERT semantic similarity for correct and incorrect responses across experimental conditions. Values are median cosine similarity (IQR). Difference is calculated as correct minus incorrect. *P* values are adjusted for multiple comparisons using the Holm method.

| Question Type | Experiment | Overall Accuracy (%) | BioMedBERT Score, Correct Responses, (IQR) | BioMedBERT Score, Incorrect Responses (IQR) | Difference | Holm-Adjusted *P* |
| --- | --- | --- | --- | --- | --- | --- |
| **ICQ** | Baseline: Full question stem with images | 59.13 (58.21-60.08) | 0.9923 (0.9901–0.9940) | 0.9928 (0.9908–0.9941) | −0.00049 | **<0.001** |
|  | W/O<sub>images</sub>: Without images | 59.13 (58.18-60.11) | 0.9921 (0.9899–0.9938) | 0.9919 (0.9898–0.9935) | +0.00022 | 0.09 |
|  | W/O<sub>stem</sub>: Without stem | 49.05 (48.07-50.00) | 0.9918 (0.9894–0.9936) | 0.9918 (0.9896–0.9933) | −0.00002 | 0.67 |
|  | W/O<sub>images+stem</sub>: Without stem and without images | 45.68 (44.70 – 46.69) | 0.9907 (0.9884–0.9928) | 0.9905 (0.9879–0.9925) | +0.00019 | **0.02** |
|  | W/O<sub>images+stem+question</sub>: Answers only | 36.49 (35.51–37.44) | 0.9900 (0.9871–0.9923) | 0.9888 (0.9855–0.9911) | +0.00117 | **<0.001** |
| **NICQ** | Baseline: Full question stem | 72.94 (72.10–73.85) | 0.9923 (0.9898–0.9940) | 0.9921 (0.9900–0.9938) | +0.00016 | 0.29 |
|  | W/O<sub>stem</sub>: Without stem | 53.53 (52.41–54.68) | 0.9912 (0.9886–0.9933) | 0.9903 (0.9878–0.9924) | +0.00092 | **<0.001** |
|  | W/O<sub>images+stem+question</sub>: Answers only | 37.36 (36.28–38.48) | 0.9899 (0.9868–0.9923) | 0.9883 (0.9849–0.9908) | +0.00162 | **<0.001** |

All *P* values are two-sided and Holm-adjusted. ICQ denotes image-containing question; IQR, interquartile range; NICQ, non-image-containing question; and W/O, without.

## DISCUSSION

LLMs have demonstrated rapidly improving performance on standardized medical exams, with recent studies reporting performance approaching or exceeding that of senior orthopaedic residents on the OITE.^5,6^ A 2023 study evaluating text-only questions from the 2020-2022 OITE reported 73.6% accuracy with GPT-4, comparable with average PGY-5 performance.^9^ More recently, Dave et al. reported 74.9% accuracy with GPT-4o on the 2024-OITE, again approximating PGY-5 performance.^6^ However, to our knowledge, no prior studies have evaluated which components of the examination question contribute to that performance. In this study, we progressively removed clinical information available to three open-source and five closed-source proprietary LLMs and evaluated their accuracy on OITE questions to quantify the performance gained by including each component.

In this study, three principal findings emerged. First, the clinical vignette was the greatest contributor to model performance, whereas removal of associated clinical images alone had minimal effect on accuracy. Second, closed-source proprietary frontier models consistently outperformed the evaluated open-source models across all experimental conditions, with larger models generally demonstrating greater accuracy within model families. Third, all models performed above the 25% accuracy expected from random chance when provided only four answer options with no clinical context or question. Together these findings suggest that clinical context is a major driver of LLM performance on the OITE. However, the above-chance accuracy observed when the question stem was removed raises the possibility that this context functions as a retrieval cue for memorized training material rather than as input for clinical reasoning.

Prior studies evaluating LLM performance on orthopaedic examinations have largely focused on overall accuracy and comparisons between models or levels of postgraduate training. For example, Rizzo et al. evaluated OpenAI GPT-3.5 and GPT-4 on OITE questions from 2020 through 2022 and examined performance by examination year, orthopedic category, presence of media, and first-versus higher order questions. They found GPT-4 outperformed GPT-3.5 across years and question categories, achieving overall accuracies ranging from 58.69% to 67.63%.^5^ Another study evaluated ChatGPT using 301 AAOS orthopaedic self-assessment questions after excluding image-containing questions. ChatGPT achieved an overall accuracy of 60.8%, with performance further stratified by subspecialty and whether questions assessed diagnosis, management, or knowledge recall.^10^ Although these studies demonstrate differences in performance according to question characteristics, they did not experimentally remove individual components of the same questions, to determine the contribution of each component to model accuracy. We used newer closed-source proprietary models from the same OpenAI family and achieved higher accuracy. There are several possibilities explaining the improved performance including better model training strategies and training data. It is plausible that the models are trained on the OITE questions given the high performance when only answer options were given to the LLMs. It is possible that the previous literature on OpenAI LLMs’ accuracy on the OITE exam overestimated the clinical reasoning ability of the models since they do not publish their training corpus, and the previous literature does not evaluate accuracy when only answers are given to the models. GPT-3.5 and GPT-4 are no longer available through the OpenAI ChatGPT service, and the models are deprecated on their API console. Given most of the previous studies used the ChatGPT service to access the models, it will be difficult to replicate their prompting strategy because it includes additional hidden context unlike when using their API console.

The contribution of imaging is difficult to establish from previous benchmarking studies. Posner et al. evaluated GPT-4 on OITE questions requiring image analysis and found significantly lower accuracy on image-requiring compared with non-image questions (47.59% vs 67.81%, *P*<0.001).^11^ In contrast, Magruder et al. found no significant difference in GPT-4 or Orthopod, a domain-specific LLM, performance between questions with and without media.^12^ Hayes et al. similarly reported comparable performance on text-only and image-associated questions from the 2019 OITE; however, image information was provided through descriptions generated by an orthopaedic specialist or a separate image-analysis system rather than interpreted directly by the multimodal LLM.^13^ These prior studies compared different groups of questions or relied on indirect representations of imaging, making it difficult to isolate the independent contribution of the image itself. Our study addresses this limitation by evaluating the same image-containing questions with and without their associated images, thereby holding the remaining question content constant. Both the open-source and closed-source families gain minimal performance by including the medical imaging and suggests multimodality remains a weakness in answering OITE questions. Additionally, because many image-containing questions require image interpretation to be answered correctly, this further raises the possibility that the models may be relying on text patterns in the clinical vignette to retrieve memorized answers rather than on interpretation of the accompanying image and answering with clinical reasoning.

The clinical vignette was the dominant contributor to model performance. Across both image-containing and non-image-containing questions, removal of the clinical vignette resulted in substantial reduction in accuracy. Clinical vignettes contained a median of 42 words for ICQs (IQR, 29-61; range, 9-129) and 41 words for NICQs (IQR, 26–57; range, 8–113). In contrast, withholding images while preserving the complete textual context resulted in only small changes in model performance, with absolute differences between the full information and image-removed conditions ranging from 0.23 to 3.23 percentage points. Model-specific paired analyses similarly demonstrated no significant effect of image removal for any evaluated model. Thus, our findings provide a potential alternative interpretation for the relatively small image-associated differences reported in prior OITE studies: for many of these questions, sufficient information to identify the correct answer may be contained within the textual clinical context. This distinction is increasingly important as multimodal LLMs are being evaluated for medical applications, where correct interpretation of radiographs and other clinical images requires models to derive clinically relevant information from the images themselves, rather than arrive at the correct answer primarily from accompanying text.

We extend previous benchmarking work by including both closed-source proprietary and open-source models. Most previous OITE investigations have centered on closed-source proprietary systems, particularly OpenAI GPT-based ones.^5,9^ Consistent with previous studies, closed-source proprietary frontier models consistently outperformed the evaluated open-source models across all experimental conditions. However, we do not know their parameter size making it difficult to determine whether performance is gained by using larger models or better LLM designs. GPT-5.6 Terra achieved the highest overall accuracy, followed by GPT-5.6 Luna and the Claude family of models, whereas the Ministral models demonstrated lower overall performance. Within the evaluated model families, larger models generally achieved higher accuracies than their smaller counterparts. Although there is a performance gap between the closed-source proprietary and open-source systems, the continued advances in open-source models may be particularly valuable for medical institutions that are seeking systems that can keep patient data secure within institutional infrastructure or require persistent model access without risk of deprecation or cloud system downtime.

An additional finding was the discordance between answer accuracy and BioMedBERT semantic reasoning similarity. Despite differences in accuracy between models, BioMedBERT similarity scores had a small range, and correct and incorrect responses demonstrated only small differences in similarity to the reference explanations. BioMedBERT similarity was also not significantly correlated with accuracy. Prior OITE benchmarking studies evaluated model-generated explanations through manual review. In those studies, researchers or orthopaedic surgeons identified reasoning patterns and errors and rated explanations for correctness, completeness, and coherence.^9,14,15^ We used BioMedBERT to provide a standardized quantitative measure of this similarity. However, the narrow range of similarity scores and the small differences between correct and incorrect responses indicate that similarity to a reference explanation does not necessarily reflect the correctness or quality of the model’s reasoning.

The most unexpected finding was the performance observed in the answer-only condition (W/O_images+stem+question_). With four answer choices and no clinical vignette, question sentence, or associated image, random selection would be expected to score an accuracy of 25%. Instead, the pooled accuracy across models remained higher across both open-source and closed-source LLMs. Although pooled accuracy across all models exceeded chance, performance differed substantially between open-source and closed-source proprietary models. The open-source Ministral models performed closer to random chance in the answer-only condition, with ICQ accuracy ranging from 25.81% to 30.41%, whereas four of the five proprietary models, Claude Sonnet 4.6, Opus 4.8, GPT-5.6 Luna, and GPT-5.6 Terra, achieved accuracies exceeding 40%, ranging from 41.47% to 45.62%. The lower performance of the Ministral models may reflect differences in their ability to infer an answer from relationships among the answer choices, or the OITE questions or related materials may have been less prevalent or absent from their training content. Because the contents of proprietary and open-source training datasets are not fully available, these potential explanations cannot be distinguished from the present results.

Interestingly, above-chance performance under their choices-only condition has also been demonstrated beyond medical examination benchmarks. Balepur et al. evaluated LLMs using multiple-choice questions spanning science, multidisciplinary academic and professional knowledge, and commonsense reasoning. When provided with only the answer choices, models performed above the majority baseline in 11 of 12 model-dataset combinations. Their analyses suggested that memorization alone did not account for this performance and identified relationships among answer choices and the ability to infer aspects of the missing question as possible contributors.^16^ Our findings extend this observation to a medical examination benchmark and suggest that answer choices themselves may provide information that contributes to model performance.

The OITE questions evaluated in this study were administered between 2020 and 2024, whereas the training-data cutoff dates for the models in this study were in 2025 and 2026 for Claude and OpenAI LLMs. Therefore, prior exposure to identical or similar content during training cannot be excluded. However, because the models’ training corpus is not public, this study cannot confirm whether there was training data leakage (i.e., we are testing on training data). Ministral models have no publicly known data cutoff dates.

Several limitations should be considered. Although three open-source model sizes were evaluated, they represented a single open-source model family, limiting the generalizability to the broader landscape of open-source LLMs. Each experimental condition used a single standardized prompting strategy; alternative prompting approaches, including explicit reasoning or chain of thought strategies, may produce different results. Third, each question was evaluated once per model and condition. Since LLM outputs can vary across repeated generations or with different hyperparameters, the present study does not evaluate the consistencies within a model’s responses. Furthermore, the OITE examinations evaluated in the study preceded the development of the tested models, the possibility of prior exposure to examination content cannot be excluded, especially for the Claude and OpenAI LLMs. Evaluation using questions created after a model’s documented training cutoff would provide a better test of LLM clinical reasoning and performance on orthopaedic exams.

Future studies should evaluate a broader range of open-source and closed-source proprietary models and examine performance on examination material both before and after documented training-data cutoffs dates. Additional evaluation using other AAOS, and orthopaedic materials would help determine whether the findings generalize beyond the OITE as a benchmark of the model’s understanding of orthopaedic surgery knowledge. The influence of prompting strategy can also be tested, including structured reasoning approaches and repeated sampling to assess response consistency. Open-source models additionally provide opportunities to update model weights using supervised fine-tuning or parameter efficient approaches, such as low rank adaptation, to improve LLM alignment. Finally, the mechanisms underlying the unexpected above-chance performance in the answer-only condition remain unclear. Future experiments could explore these and could vary the number and composition of distractor answers, repeat randomization of answer order, and introduce novel incorrect alternatives. Further experiments using novel questions and randomized answer-option order could help distinguish whether above-chance performance arises from information contained within the answer choices, prior exposure to examination content, or other features of model behavior. As clinical imaging is important in orthopaedic assessments, further work may benefit from improved multimodality. Future work should have clinicians evaluate the quality of the LLM answer explanations and review image saliency maps to improve model answer and explanation trustworthiness.

## CONCLUSION

LLMs are increasingly being used in clinical context and characterizing their clinical reasoning is important for building clinician trust. Standardized medical exams, such as the orthopaedic in-training exam, are commonly used to benchmark model performance. This study showed that the clinical context predominantly drove the improved accuracy, whereas removal of accompanying images had minimal effect. Closed-source proprietary models consistently outperformed the Ministral open-source models. However, all models maintained above-chance performance when provided only the answer options suggesting the possibility that the models may have been trained on the OITE questions. BioMedBERT indicated that model-generated explanations were similarly aligned in meaning with the reference OITE explanations across models, despite substantial differences in answer accuracy, and did not significantly correlate with accuracy. Collectively, these findings suggest that reporting performance on older standardized medical examinations may be overestimating clinical reasoning ability and highlight the importance of evaluating how LLMs use the information provided when assessing their potential applications in medical education and clinical practice.

## Data Availability

The OITE examination questions and answer keys analyzed in this study are proprietary materials of the American Academy of Orthopaedic Surgeons and cannot be redistributed by the authors. These materials may be obtained from the American Academy of Orthopaedic Surgeons subject to its applicable access requirements and terms of use.

## REFERENCES

1. Kung TH, Cheatham M, Medenilla A, et al. Performance of ChatGPT on USMLE: Potential for AI-assisted medical education using large language models. PLOS Digital Health [Internet] 2023 [cited 2026 Aug 10];2(2):e0000198. Available from: https://journals.plos.org/digitalhealth/article?id=10.1371/journal.pdig.0000198

2. Safranek CW, Sidamon-Eristoff AE, Gilson A, Chartash D. The Role of Large Language Models in Medical Education: Applications and Implications. JMIR Med Educ [Internet] 2023 [cited 2026 Aug 10];9:e50945. Available from: https://mededu.jmir.org/2023/1/e50945

3. Lubitz M, Latario L. Performance of Two Artificial Intelligence Generative Language Models on the Orthopaedic In-Training Examination. Orthopedics [Internet] 2024 [cited 2026 Aug 10];47(3):e146–50. Available from: https://journals.healio.com/doi/10.3928/01477447-20240304-02

4. Xu AY, Singh M, Balmaceno-Criss M, et al. Comparitive performance of artificial intelligence-based large language models on the orthopedic in-training examination. J Orthop Surg (Hong Kong) 2025;33(1):10225536241268789.

5. Rizzo MG, Cai N, Constantinescu D. The performance of ChatGPT on orthopaedic in-service training exams: A comparative study of the GPT-3.5 turbo and GPT-4 models in orthopaedic education. Journal of Orthopaedics [Internet] 2024 [cited 2026 Aug 10];50:70–5. Available from: https://www.sciencedirect.com/science/article/pii/S0972978X2300332X

6. Dave R, Vediya N, Sharma N, Shah A, Whelan D, Wolfstadt J. Large Language Models Outperform PGY-5 Residents on the Orthopaedic In-Training Examination: A Comparative Analysis of Six Cutting-Edge Large Language Models. J Am Acad Orthop Surg [Internet] 2026 [cited 2026 Aug 10];34(13):e1771–80. Available from: https://journals.lww.com/10.5435/JAAOS-D-25-01242

7. Ko S, Lee J, Ko K, Kim J. Benchmarking Open-Source Vision Language Models in Orthopedic In-Training Examination: A Comparison with Residents, Domain-Specific Evaluation, and Parameter Scaling. Clinics in Orthopedic Surgery [Internet] 2026 [cited 2026 Aug 10];18(1):159–66. Available from: 10.4055/cios25183

8. Gu Y, Tinn R, Cheng H, et al. Domain-Specific Language Model Pretraining for Biomedical Natural Language Processing. ACM Trans Comput Healthcare [Internet] 2022 [cited 2026 Aug 30];3(1):1–23. Available from: http://arxiv.org/abs/2007.15779

9. Kung JE, Marshall C, Gauthier C, Gonzalez TA, Jackson JB. Evaluating ChatGPT Performance on the Orthopaedic In-Training Examination. JB JS Open Access [Internet] 2023 [cited 2026 Aug 10];8(3):e23.00056. Available from: https://pmc.ncbi.nlm.nih.gov/articles/PMC10484364/

10. Isleem UN, Zaidat B, Ren R, et al. Can generative artificial intelligence pass the orthopaedic board examination? J Orthop [Internet] 2023 [cited 2026 Aug 10];53:27–33. Available from: https://pmc.ncbi.nlm.nih.gov/articles/PMC10912220/

11. Posner KM, Bakus C, Basralian G, et al. Evaluating ChatGPT’s Capabilities on Orthopedic Training Examinations: An Analysis of New Image Processing Features. Cureus [Internet] 2024 [cited 2026 Aug 10];16. Available from: https://cureus.com/articles/228626-evaluating-chatgpts-capabilities-on-orthopedic-training-examinations-an-analysis-of-new-image-processing-features

12. Magruder ML, Miskiewicz M, Rodriguez AN, Ng M, Abdelgawad A. Comparison of ChatGPT plus (version 4.0) and pretrained AI model (Orthopod) on orthopaedic in-training exam (OITE). The Surgeon [Internet] 2025 [cited 2026 Aug 10];23(3):187–91. Available from: https://www.sciencedirect.com/science/article/pii/S1479666X2500054X

13. Hayes DS, Foster BK, Makar G, et al. Artificial Intelligence in Orthopaedics: Performance of ChatGPT on Text and Image Questions on a Complete AAOS Orthopaedic In-Training Examination (OITE). Journal of Surgical Education [Internet] 2024 [cited 2026 Aug 10];81(11):1645–9. Available from: https://www.sciencedirect.com/science/article/pii/S1931720424003799

14. Mendiratta D, Herzog I, Singh R, et al. Utility of ChatGPT as a preparation tool for the Orthopaedic In-Training Examination. Journal of Experimental Orthopaedics [Internet] 2025 [cited 2026 Aug 10];12(1):e70135. Available from: https://onlinelibrary.wiley.com/doi/abs/10.1002/jeo2.70135

15. Balepur N, Ravichander A, Rudinger R. Artifacts or Abduction: How Do LLMs Answer Multiple-Choice Questions Without the Question? [Internet]. 2024 [cited 2026 Aug 10];Available from: http://arxiv.org/abs/2402.12483

