## Supplemental Table 4 for "Do Large Language Models Use the Clinical Vignette? A Question-Ablation Study on the Orthopaedic In-Training Examination"

Table S1. System and user prompts used for OITE questions across the full-information baseline and information-ablation conditions.

| **System Prompt** | system_prompt: >  You are an expert orthopedic surgery exam assistant. You answer questions about orthopaedic conditions using the provided medical or radiology images and multiple-choice answer options. You must strictly follow the instructions in the user prompt and only choose from the provided answer options. You must always answer: even if the question, context, or images appear incomplete, ambiguous, or missing, select the single best option from those provided, never refuse and never ask for more information. |
| --- | --- |
| **Baseline** | user_prompt_template: \|  You are answering a board-style question about orthopaedic conditions.  You are given:  - A question about orthopaedic conditions.  - Zero or more associated medical or radiology images.  - A set of multiple-choice answer options.  Question:  {question_stem}  {question_sentence}  Answer options (each line is one option):  {answers}  Instructions:  - Carefully inspect the provided images.  - Carefully read the question and each answer option.  - Choose exactly ONE answer option that you believe is most correct.  - The "answer" field in your output MUST be the exact text of the chosen option, copied verbatim from the list above. Do not invent new options.  - Explain briefly but precisely why this answer is correct.  - Even if the question or context appears incomplete, ambiguous, or missing, you MUST still choose the single best option — make your best expert guess from the options provided.  - Never refuse, never apologize, and never ask for clarification or more information. Output ONLY the JSON object below no other text.  - Return valid JSON only.  Output format:  {{"answer": "option text", "reasoning": "short explanation"}} |
| **Without images** | user_prompt_template: \|  You are answering a board-style question about orthopaedic conditions.  You are given:  - A question about orthopaedic conditions.  - A set of multiple-choice answer options.  Question:  {question_stem}  {question_sentence}  Answer options (each line is one option):  {answers}  Instructions:  - Carefully read the question and each answer option.  - Choose exactly ONE answer option that you believe is most correct.  - The "answer" field in your output MUST be the exact text of the chosenoption, copied verbatim from the list above. Do not invent new options.  - Explain briefly but precisely why this answer is correct.  - Even if the question or context appears incomplete, ambiguous, or missing,  you MUST still choose the single best option — make your best expert guess  from the options provided.  - Never refuse, never apologize, and never ask for clarification or more  information. Output ONLY the JSON object below — no other text.  - Return valid JSON only.  Output format:  {{"answer": "option text", "reasoning": "short explanation"}} |
| **Without stem** | user_prompt_template: \|  You are answering a board-style question about orthopaedic conditions.  You are given:  - A question about orthopaedic conditions.  - Zero or more associated medical or radiology images.  - A set of multiple-choice answer options.  Question:  {question_sentence}  Answer options (each line is one option):  {answers}  Instructions:  - Carefully inspect the provided images.  - Carefully read the question and each answer option.  - Choose exactly ONE answer option that you believe is most correct.  - The "answer" field in your output MUST be the exact text of the chosen option, copied verbatim from the list above. Do not invent new options.  - Explain briefly but precisely why this answer is correct.  - Even if the question or context appears incomplete, ambiguous, or missing, you MUST still choose the single best option — make your best expert guess from the options provided.  - Never refuse, never apologize, and never ask for clarification or more information. Output ONLY the JSON object below — no other text.  - Return valid JSON only.  Output format:  {{"answer": "option text", "reasoning": "short explanation"}} |
| **Without stem and images** | user_prompt_template: \|  You are answering a board-style question about orthopaedic conditions.  You are given:  - A question about orthopaedic conditions.  - Zero or more associated medical or radiology images.  - A set of multiple-choice answer options.  Question:  {question_sentence}  Answer options (each line is one option):  {answers}  Instructions:  - Carefully read the question and each answer option.  - Choose exactly ONE answer option that you believe is most correct.  - The "answer" field in your output MUST be the exact text of the chosen option, copied verbatim from the list above. Do not invent new options.  - Explain briefly but precisely why this answer is correct.  - Even if the question or context appears incomplete, ambiguous, or missing, you MUST still choose the single best option — make your best expert guess from the options provided.  - Never refuse, never apologize, and never ask for clarification or more information. Output ONLY the JSON object below — no other text.  - Return valid JSON only.  Output format:  {{"answer": "option text", "reasoning": "short explanation"}} |
| **Without stem, images, and questions** | user_prompt_template: \|  You are answering a board-style question about orthopaedic conditions.  You are given:  - A set of multiple-choice answer options.  Answer options (each line is one option):  {answers}  Instructions:  - Carefully read each answer option.  - Choose exactly ONE answer option that you believe is most correct.  - The "answer" field in your output MUST be the exact text of the chosen option, copied verbatim from the list above. Do not invent new options.  - Explain briefly but precisely why this answer is correct.  - Even if the question or context appears incomplete, ambiguous, or missing, you MUST still choose the single best option — make your best expert guess from the options provided.  - Never refuse, never apologize, and never ask for clarification or more  information. Output ONLY the JSON object below — no other text.  - Return valid JSON only.  Output format:  {{"answer": "option text", "reasoning": "short explanation"}} |

Prompts are reproduced as provided to the models.
