## Supplemental Table 4 for "Do Large Language Models Use the Clinical Vignette? A Question-Ablation Study on the Orthopaedic In-Training Examination"

Table S2. Model-specific paired comparisons of accuracy across experimental conditions. ICQs: questions containing clinical images.

| **Model** | **Comparison (A vs B)** | **Accuracy A (%)** | **Accuracy B (%)** | **Difference in Accuracy (%)** | **McNemar adjusted *P*** |
| --- | --- | --- | --- | --- | --- |
| Ministral-3 3B | Baseline vs W/O_stem_ | 32.72 (31.34–34.33) | 29.95 (28.34–31.57) | 2.76 | 1.000 |
|  | Baseline vs W/O_images+stem_ | 32.72 (31.34–34.33) | 27.42 (25.81–28.80) | 5.30 | 0.24 |
|  | Baseline vs W/O_images_ | 32.72 (31.34–34.33) | 30.65 (29.26–32.26) | 2.07 | 1.000 |
|  | Baseline vs W/O_images+stem+question_ | 32.72 (31.34–34.33) | 25.81 (24.42–27.19) | 6.91 | 0.09 |
|  | W/O_stem_ vs W/O_images+stem_ | 29.95 (28.34–31.57) | 27.42 (25.81–28.80) | 2.53 | 1.00 |
| Ministral-3 8B | Baseline vs W/O_stem_ | 38.71 (37.10–40.32) | 29.95 (28.57–31.57) | 8.76 | **0.001** |
|  | Baseline vs W/O_images+stem_ | 38.71 (37.10–40.32) | 29.72 (28.34–31.11) | 8.99 | **0.002** |
|  | Baseline vs W/O_images_ | 38.71 (37.10–40.32) | 38.25 (36.64–39.86) | 0.46 | 1.000 |
|  | Baseline vs W/O_images+stem+question_ | 38.71 (37.10–40.32) | 27.88 (26.27–29.26) | 10.83 | **<0.001** |
|  | W/O_stem_ vs W/O_images+stem_ | 29.95 (28.57–31.57) | 29.72 (28.34–31.11) | 0.23 | 1.000 |
| Ministral-3 14B | Baseline vs W/O_stem_ | 40.09 (38.25–41.71) | 37.56 (35.94–39.17) | 2.53 | 0.36 |
|  | Baseline vs W/O_images+stem_ | 40.09 (38.25–41.71) | 33.64 (32.03–35.25) | 6.45 | 0.100 |
|  | Baseline vs W/O_images_ | 40.09 (38.25–41.71) | 42.86 (41.24–44.47) | −2.76 | 0.22 |
|  | Baseline vs W/O_images+stem+question_ | 40.09 (38.25–41.71) | 30.41 (28.80–32.03) | 9.68 | **0.004** |
|  | W/O_stem_ vs W/O_images+stem_ | 37.56 (35.94–39.17) | 33.64 (32.03–35.25) | 3.92 | 0.22 |
| Claude Haiku 4.5 | Baseline vs W/O_stem_ | 52.53 (51.15–53.92) | 41.94 (40.32–43.32) | 10.60 | **<0.001** |
|  | Baseline vs W/O_images+stem_ | 52.53 (51.15–53.92) | 42.17 (40.78–43.55) | 10.37 | **<0.001** |
|  | Baseline vs W/O_images_ | 52.53 (51.15–53.92) | 53.69 (52.07–55.30) | −1.15 | 1.00 |
|  | Baseline vs W/O_images+stem+question_ | 52.53 (51.15–53.92) | 32.03 (30.65–33.41) | 20.51 | **<0.001** |
|  | W/O_stem_ vs W/O_images+stem_ | 41.94 (40.32–43.32) | 42.17 (40.78–43.55) | −0.23 | 1.00 |
| Claude Sonnet 4.6 | Baseline vs W/O_stem_ | 70.97 (69.53–72.58) | 55.53 (53.92–57.14) | 15.44 | **<0.001** |
|  | Baseline vs W/O_images+stem_ | 70.97 (69.53–72.58) | 53.23 (51.84–55.07) | 17.74 | **<0.001** |
|  | Baseline vs W/O_images_ | 70.97 (69.53–72.58) | 72.58 (71.20–73.96) | −1.61 | 0.71 |
|  | Baseline vs W/O_images+stem+question_ | 70.97 (69.53–72.58) | 43.32 (41.94–44.93) | 27.65 | **<0.001** |
|  | W/O_stem_ vs W/O_images+stem_ | 55.53 (53.92–57.14) | 53.23 (51.84–55.07) | 2.30 | 0.71 |
| Claude Opus 4.8 | Baseline vs W/O_stem_ | 78.80 (77.42–80.18) | 64.06 (62.62–65.67) | 14.75 | **<0.001** |
|  | Baseline vs W/O_images+stem_ | 78.80 (77.42–80.18) | 56.91 (55.30–58.53) | 21.89 | **<0.001** |
|  | Baseline vs W/O_images_ | 78.80 (77.42–80.18) | 79.26 (77.88–80.65) | −0.46 | 0.88 |
|  | Baseline vs W/O_images+stem+question_ | 78.80 (77.42–80.18) | 45.62 (44.24–47.24) | 33.18 | **<0.001** |
|  | W/O_stem_ vs W/O_images+stem_ | 64.06 (62.62–65.67) | 56.91 (55.30–58.53) | 7.14 | **0.02** |
| GPT-5.6 Luna | Baseline vs W/O_stem_ | 77.42 (76.04–78.80) | 65.21 (63.59–66.82) | 12.21 | **<0.001** |
|  | Baseline vs W/O_images+stem_ | 77.42 (76.04–78.80) | 58.76 (57.37–60.60) | 18.66 | **<0.001** |
|  | Baseline vs W/O_images_ | 77.42 (76.04–78.80) | 74.19 (72.81–75.58) | 3.23 | 0.07 |
|  | Baseline vs W/O_images+stem+question_ | 77.42 (76.04–78.80) | 41.47 (39.86–43.09) | 35.94 | **<0.001** |
|  | W/O_stem_ vs W/O_images+stem_ | 65.21 (63.59–66.82) | 58.76 (57.37–60.60) | 6.45 | **0.02** |
| GPT-5.6 Terra | Baseline vs W/Ostem | 81.80 (80.41–82.95) | 68.20 (66.59–69.59) | 13.59 | **<0.001** |
|  | Baseline vs W/O_images+stem_ | 81.80 (80.41–82.95) | 63.59 (62.21–65.21) | 18.20 | **<0.001** |
|  | Baseline vs W/O_images_ | 81.80 (80.41–82.95) | 81.57 (80.41–82.95) | 0.23 | 1.000 |
|  | Baseline vs W/O_images+stem+question_ | 81.80 (80.41–82.95) | 45.39 (43.78–47.00) | 36.41 | **<0.001** |
|  | W/O_stem_ vs W/O_images+stem_ | 68.20 (66.59–69.59) | 63.59 (62.21–65.21) | 4.61 | 0.17 |

Differences are calculated as condition A minus condition B and are reported in percentage points. P values are from two-sided McNemar tests with Holm adjustment. ICQ denotes image-containing question; IQR, interquartile range; and W/O, without.
