## Supplemental Table 4 for "Do Large Language Models Use the Clinical Vignette? A Question-Ablation Study on the Orthopaedic In-Training Examination"

Table S3. Model-specific paired comparisons of accuracy across experimental conditions. NICQs: questions without clinical images.

| **Model** | **Comparison** | **Accuracy A (%)** | | **Accuracy B (%)** | **Difference in Accuracy (%)** | **McNemar adjusted *P*** |
| --- | --- | --- | --- | --- | --- | --- |
| Ministral-3 3B | Baseline vs W/O_stem_ | 41.06 (39.39–42.74) | 34.08 (32.40–36.03) | | 6.98 | **0.010** |
|  | Baseline vs W/O_stem+question_ | 41.06 (39.39–42.74) | 27.09 (25.42–29.05) | | 13.97 | **<0.001** |
|  | W/O_stem_ vs W/O_stem+question_ | 34.08 (32.40–36.03) | 27.09 (25.42–29.05) | | 6.98 | **0.010** |
| Ministral-3 8B | Baseline vs W/O_stem_ | 54.75 (53.07–56.70) | 41.06 (39.39–42.74) | | 13.69 | **<0.001** |
|  | Baseline vs W/O_stem+question_ | 54.75 (53.07–56.70) | 31.01 (29.33–32.68) | | 23.74 | **<0.001** |
|  | W/O_stem_ vs W/O_stem+question_ | 41.06 (39.39–42.74) | 31.01 (29.33–32.68) | | 10.06 | **<0.001** |
| Ministral-3 14B | Baseline vs W/O_stem_ | 53.91 (51.96–55.87) | 40.78 (39.11–42.74) | | 13.13 | **<0.001** |
|  | Baseline vs W/O_stem+question_ | 53.91 (51.96–55.87) | 29.05 (27.65–30.73) | | 24.86 | **<0.001** |
|  | W/O_stem_ vs W/O_stem+question_ | 40.78 (39.11–42.74) | 29.05 (27.65–30.73) | | 11.73 | **<0.001** |
| Claude Haiku 4.5 | Baseline vs W/O_stem_ | 70.67 (68.99–72.35) | 48.32 (46.65–50.28) | | 22.35 | **<0.001** |
|  | Baseline vs W/O_stem+question_ | 70.67 (68.99–72.35) | 32.12 (30.45–33.80) | | 38.55 | **<0.001** |
|  | W/O_stem_ vs W/O_stem+question_ | 48.32 (46.65–50.28) | 32.12 (30.45–33.80) | | 16.20 | **<0.001** |
| Claude Sonnet 4.6 | Baseline vs W/O_stem_ | 87.15 (86.03–88.55) | 62.29 (60.34–63.97) | | 24.86 | **<0.001** |
|  | Baseline vs W/O_stem+question_ | 87.15 (86.03–88.55) | 44.97 (43.30–46.65) | | 42.18 | **<0.001** |
|  | W/O_stem_ vs W/O_stem+question_ | 62.29 (60.34–63.97) | 44.97 (43.30–46.65) | | 17.32 | **<0.001** |
| Claude Opus 4.8 | Baseline vs W/O_stem_ | 89.94 (88.83–91.06) | 65.92 (64.25–67.60) | | 24.02 | **<0.001** |
|  | Baseline vs W/O_stem+question_ | 89.94 (88.83–91.06) | 45.25 (43.30–46.93) | | 44.69 | **<0.001** |
|  | W/O_stem_ vs W/O_stem+question_ | 65.92 (64.25–67.60) | 45.25 (43.30–46.93) | | 20.67 | **<0.001** |
| GPT-5.6 Luna | Baseline vs W/O_stem_ | 91.90 (90.78–92.74) | 66.76 (65.08–68.44) | | 25.14 | **<0.001** |
|  | Baseline vs W/O_stem+question_ | 91.90 (90.78–92.74) | 43.30 (41.34–44.97) | | 48.60 | **<0.001** |
|  | W/O_stem_ vs W/O_stem+question_ | 66.76 (65.08–68.44) | 43.30 (41.34–44.97) | | 23.46 | **<0.001** |
| GPT-5.6 Terra | Baseline vs W/O_stem_ | 94.13 (93.30–94.97) | 68.99 (67.60–70.67) | | 25.14 | **<0.001** |
|  | Baseline vs W/O_stem+question_ | 94.13 (93.30–94.97) | 46.09 (44.41–47.77) | | 48.04 | **<0.001** |
|  | W/O_stem_ vs W/O_stem+question_ | 68.99 (67.60–70.67) | 46.09 (44.41–47.77) | | 22.91 | **<0.001** |

Differences are calculated as condition A minus condition B and are reported in percentage points. *P* values are from two-sided McNemar tests with Holm adjustment. IQR denotes interquartile range; NICQ, non–image-containing question; and W/O, without.
