## Supplemental Table 4 for "Do Large Language Models Use the Clinical Vignette? A Question-Ablation Study on the Orthopaedic In-Training Examination"

Table S4. Model-specific accuracy and BioMedBERT semantic similarity for image-containing and non-image containing OITE questions under the full-information condition (Baseline)

| **Model** | **ICQs** | | **NICQs** | |
| --- | --- | --- | --- | --- |
|  | **Accuracy (%)** | **BioMedBERT Similarity** | **Accuracy (%)** | **BioMedBERT Similarity** |
| Ministral-3 3B | 32.72% (31.34–34.33) | 0.9927 | 41.06% (39.39–42.74) | 0.9918 |
| Ministral-3 8B | 38.71% (37.10–40.32) | 0.9930 | 54.75% (53.07–56.70) | 0.9925 |
| Ministral-3 14B | 40.09% (38.25–41.71) | 0.9932 | 53.91% (51.96–55.87) | 0.9908 |
| Claude Haiku 4.5 | 52.53% (51.15–53.92) | 0.9921 | 70.67% (68.99–72.35) | 0.9931 |
| Claude Sonnet 4.6 | 70.97% (69.53–72.58) | 0.9931 | 87.15% (86.03–88.55) | 0.9935 |
| Claude Opus 4.8 | 78.80% (77.42–80.18) | 0.9927 | 89.94% (88.83–91.06) | 0.9922 |
| GPT-5.6 Luna | 77.42% (76.04–78.80) | 0.9893 | 91.90% (90.78–92.74) | 0.9887 |
| GPT-5.6 Terra | 81.80% (80.41–82.95) | 0.9894 | 94.13% (93.30–94.97) | 0.9888 |

BioMedBERT values are reported as median cosine similarity with IQR. ICQ denotes image-containing question; IQR, interquartile range; and NICQ, non–image-containing question.
